# Machine Learning Generalizability Across Healthcare Settings: Insights from multi-site COVID-19 screening

**DOI:** 10.1101/2022.02.09.22269744

**Authors:** Jenny Yang, Andrew A. S. Soltan, David A. Clifton

## Abstract

As patient health information is highly regulated due to privacy concerns, the majority of machine learning (ML)-based healthcare studies are unable to test on external patient cohorts, resulting in a gap between locally reported model performance and cross-site generalizability. Different approaches have been introduced for developing models across multiple clinical sites, however no studies have compared methods for translating ready-made models for adoption in new settings. We introduce three methods to do this – (1) applying a ready-made model “as-is”; (2) readjusting the decision threshold on the output of a ready-made model using site-specific data; and (3) finetuning a ready-made model using site-specific data via transfer learning. Using a case study of COVID-19 diagnosis across four NHS Hospital Trusts, we show that all methods achieve clinically-effective performances (NPV >0.959), with transfer learning achieving the best results (mean AUROCs between 0.870-0.925). Our models demonstrate that site-specific customization improves predictive performance when compared to other ready-made approaches.

## Introduction

The advantages of applying machine learning (ML) methods within healthcare have become increasingly evident, particularly for the purposes of predictive modelling. As the development of ML-based models continues to surge, greater attention is being given to a model’s reproducibility. This is especially critical within healthcare, as algorithmic findings can directly influence clinical decision-making and patient care.

In ML, different types of reproducibility have been commonly discussed, including technical, statistical, and conceptual reproducibility (Gunderson & Kjensmo, 2018; McDermott et al., 2021). Technical (or methods) reproducibility refers to implementing computational procedures as precisely as possible (using the same code, dataset, etc.) to yield the same results as those reported; statistical reproducibility refers to upholding statistically similar results under resampled conditions (such as using different subsamples of data for training); and conceptual (or results) reproducibility refers to obtaining similar findings under new conditions that match the theoretical description of the original experiment (Gunderson & Kjensmo, 2018; McDermott et al., 2021). In this study, we focus on addressing conceptual reproducibility, specifically in the context of external validity and generalizability.

For predictive modelling of clinical tasks, we define generalizability as the ability of a model to perform its purpose on data from a new, independent cohort of patients (i.e., patients whose data were never used during the development process), akin to the external validation of a model (Azad et al., 2021). If a model does not satisfy this definition, then it may not have learned the true relationship between the features and the target outcome, but instead, has learned consequent biases from data generation and processing protocols. This makes it harder both to reproduce results when deployed in practice, as well as produce accurate results in general. One study recently found that only ≈ 23% of ML-based healthcare papers used multiple datasets (McDermott et al., 2021). This is a critical issue, as different healthcare settings can vary in terms of unobserved confounders, protocols, deployment environments, and drift over time, making it challenging for cross-centre generalizability. Thus, validating a model on only one dataset – particularly, one from the same cohort as the training data – is discouraged, as researchers have found large performance gaps when testing on external patient cohorts (Barak-Corren et al., 2021; Burns et al., 2020; McDermott et al., 2021). Although encouraged, validating on more datasets than the original development dataset is not always feasible, as there may be ethical, technical, or financial challenges in terms of sharing clinical data (Figueiredo, 2017; Malin & Goodman, 2018). Thus, to make ML healthcare models more accessible across different hospital settings, different model-customization approaches are needed to translate site-specific, locally-trained models to new settings.

Additionally, training clinically-effective models is not a trivial task – it can be hard to collect large, diverse, and curated datasets for training; there can be thousands of features, and models can be large and complex. If there is not enough data, then models will be unable to generalize with new, unseen data; and if there are large numbers of features or the model being trained is complex, large computational resources may be required to reach convergence (i.e., completion of the training process). These reasons can make it impractical for some sites to train models completely from scratch, further highlighting the importance of effectively using ready-made models.

In this study, we outline and evaluate three different frameworks for adopting single-site, ready-made ML models for use in new, independent hospital settings, which are as follows: (1) applying a ready-made model “as-is”; (2) readjusting the decision threshold on the output of a ready-made model using site-specific data; and (3) finetuning a ready-made model using site-specific data, by means of transfer learning. In the latter, a subset of the large, complex model is retrained, or finetuned, on the new target dataset – without retraining the entire model. We compare these methods to a combined-site approach, in which a model is trained with data from multiple sites, as well as comparing to models trained solely on data from the same site at which it will be used (i.e., an internally-validated model using data from the same site). Each of these methods can be applied under different circumstances, which are detailed in the “Methods” section. We evaluated these various approaches in the context of rapid COVID-19 diagnosis, using real-world clinical data from emergency admissions to four independent UK National Health Service (NHS) Trusts.

We aim to build on a previous study introduced by Soltan et al. (2022), where an ML pipeline (based on XGBoost) was used to screen patients attending hospital emergency departments for COVID-19. In this study, the authors trained and tested models using data from one NHS trust (Oxford University Hospitals; OUH), thereafter externally and prospectively validating the models across four independent trusts. Their results showed that their pipeline performed effectively as a screening test for COVID-19 across multiple sites. Although external validation sets were used, these datasets were preprocessed during model development and validation using the same processing protocols used with the original training data. For example, the imputation method used for all sites was determined using the training data and subsequently applied to each of the respective three test sets. This latter approach results in good performance, as the test sets have been processed using techniques based on the data distributions of the training set – this avoids difficulties that might otherwise arise if preprocessing techniques vary between sites. This format of external validation requires either (i) that data from different sites are all available in one place, (ii) that data from the original training location is available at the new test sites, or (iii) that the original preprocessing pipeline used (including normalization and imputation scalars) can be accessed by new test sites. However, in many cases, different sites may not have any of these three forms of access. Thus, using the same datasets as Soltan et al. (2022), we developed and tested our models such that data from different sites are processed completely independent of the training data – as would be required for many external validation exercises in practice. To investigate a role for transfer learning, the current work uses a complex neural network approach which was previously shown to be effective at screening for COVID-19 (Yang et al., 2022), instead of the previously described XGBoost approach. Tree-based algorithms, such as XGBoost, depend on the availability of the entire dataset, such that transfer learning is not typically feasible.

There have been different approaches used to address model generalization for clinical questions, including: 1) combined-site training, where training data comes from multiple sites (Grist et al., 2020; Ihlen et al., 2020; Nunes et al., 2020; Zeng et al., 2018); and 2) federated-learning, whereby a model is collaboratively trained at different sites through the sharing of model parameters but not the data itself (Bai et al., 2021; Dayan et al., 2021). These techniques require that data from multiple sites are available at the same time for model training, and that different institutions have agreed to have their data used for a centralized task, at the same time. This can be laborious to implement, as different institutions can have different data collection, recording, and sharing protocols, as well as different technical requirements. Additionally, such techniques are based on multi-site training, rather than the application of ready-made models in new environments.

Recently, Barack-Corren et al. (2021) compared different implementation strategies for developing prediction models at different hospital sites. Using an Emergency Department (ED) disposition case study, they applied their prediction model to three different hospital sites. They evaluated a ready-made approach, where they developed a model at one site then applied it “as-is” to data from the other sites; and a “calibrated” approach, where they trained a model at one site, then selected the top twenty features to be used for model training at the other sites. They also compared these methods to a combined-site approach (one model is trained on data from multiple sites), as well as a single-site baseline (separate models are developed and validated independently). The “calibrated” approach described still requires site-specific training at the new site. It does not actually apply a ready-made model to new data, but instead allows the new location to bypass the variable-selection process by providing the list of features to be used in training. Although this may save time, some features may be site-specific, and thus, may not be available at the new site. Additionally, the new site still needs to completely train a model from scratch. In terms of their combined-site model, they combined data from all sites into one centralized dataset, before implementing their previously developed admissions-prediction framework (Barack-Corren et al., 2017). Here, they normalized all vital sign features by age; however, did not describe if the training, validation, and test sets were transformed using the mean and standard deviation of the entire dataset, just the training set, or each respective test set independently. If using either of the first two techniques, then the test sets have already been biased using transforms based on the training set’s distribution. This results in performance appearing better at the sites that participated in the training, but not at new sites or new data from the same sites that aren’t processed with the same transforms.

Furthermore, the sharing of code and data is widely seen as being crucial to reproducibility; however, in many medical fields, they are rarely made available, as patient health information is highly regulated due to privacy concerns (Haibe-Kains et al., 2020). Thus, we built and tested our models under the assumption that external sites do not have access to the original data used to train a model, nor any transforms used during preprocessing (including normalization, imputation, or standardization transforms), as these are fitted on the training data and can be considered sensitive.

To summarize, the main contributions of this paper are as follows:

- We propose three methods for adopting single-site, ready-made ML models for use in new, independent hospital settings.
- We provide the first demonstration and comparison of cross-site implementation strategies for ready-made ML-based healthcare models, through a case study of COVID-19 diagnosis.

## Methods

We trained neural network models to predict the COVID-19 status of patients presenting to hospital emergency departments across four independent UK National Health Service (NHS) Trusts – Oxford University Hospitals NHS Foundation Trust (OUH), University Hospitals Birmingham NHS Trust (UHB), Bedfordshire Hospitals NHS Foundations Trust (BH), and Portsmouth Hospitals University NHS Trust (PUH). The inclusion and exclusion criterion used can be found in Appendix A of the Supplementary Material.

For each site, we split the data into training, validation, and test sets. From OUH, we had two data extracts. The first data extract consisted of 114,957 COVID-free patient presentations prior to the global COVID-19 outbreak, and 701 COVID-positive (by PCR testing) patient presentations during the first wave of the COVID-19 epidemic in the UK (pre-pandemic and UK “wave one” cases, to June 30, 2020). We split this cohort into a training and validation set using an 80:20 ratio, respectively. This ensures that the label of COVID-19 status is correct during training. Our second data extract consisted of 20,845 COVID-negative patients and 2,012 COVID-positive patients (PCR-confirmed) and was used as the test set (presentations from UK “wave two”, from October 1, 2020 to March 6, 2021). For PUH, UHB, and BH, we had only one extract each, consisting exclusively of presentations from during the COVID-19 outbreak, and thus used a 60:20:20 split for the training, validation, and test sets, respectively. A summary of population statistics can be found in Table 1, and a summary of training, validation, and test cohorts can be found in Table 2.

**Table 1:**
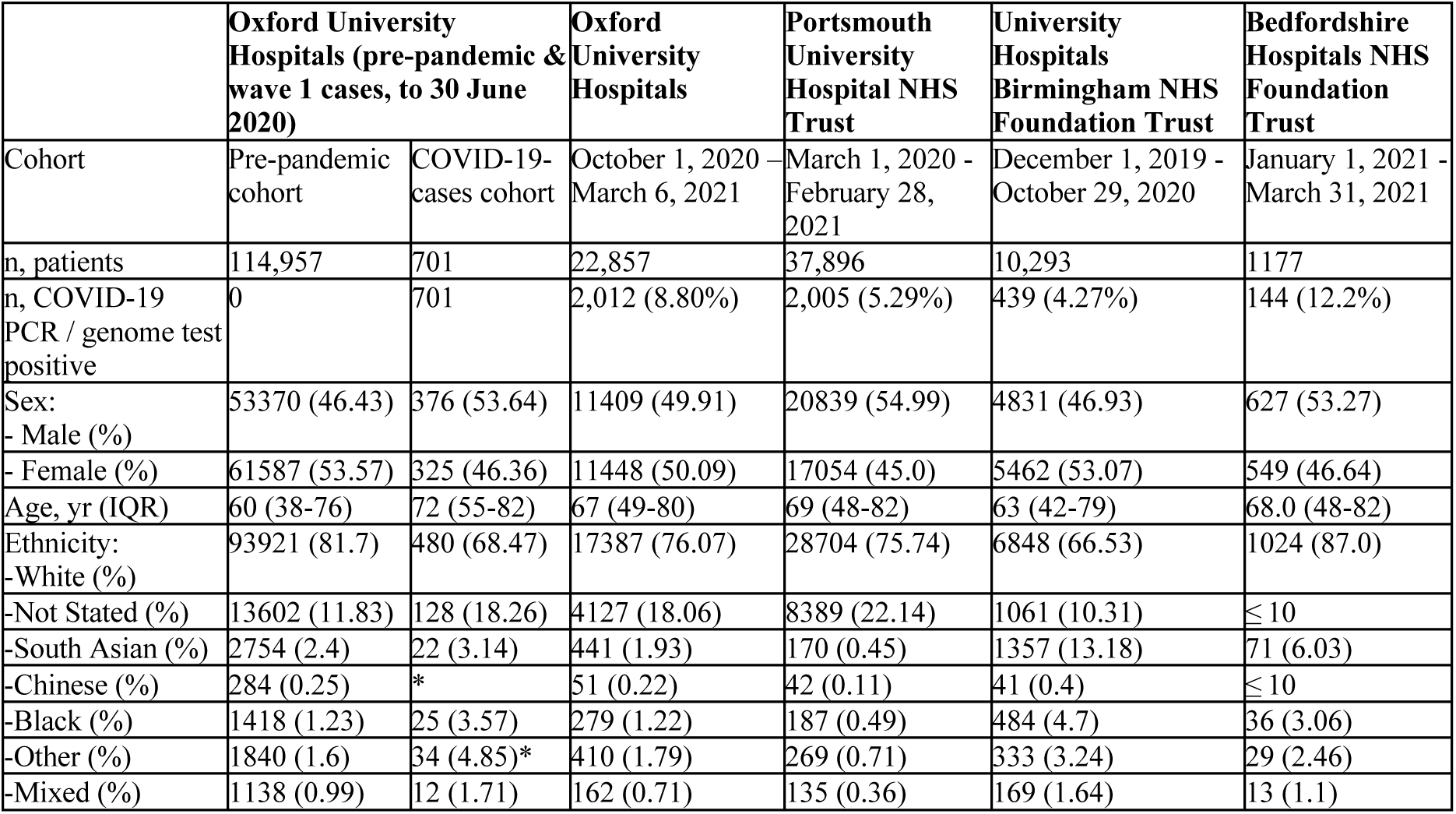
Summary population characteristics for OUH, PUH, UHB, and BH cohorts. *indicates merging for statistical disclosure control.

**Table 2:** Training, validation, and test set distributions.

|  | Oxford University Hospitals (pre-pandemic & wave 1 cases, to 30 June 2020) | Oxford University Hospitals | Portsmouth University Hospital NHS Trust | University Hospitals Birmingham NHS Foundation Trust | Bedfordshire Hospitals NHS Foundation Trust |
| --- | --- | --- | --- | --- | --- |
| Training Set | 91,970 (negative)<br>556 (positive) |  | 21,555 (negative)<br>1,182 (positive) | 5,920 (negative)<br>255 (positive) | 620 (negative)<br>85 (positive) |
| Validation Set | 22,987 (negative)<br>145 (positive) |  | 7,140 (negative)<br>439 (positive) | 1,966 (negative)<br>93 (positive) | 206 (negative)<br>30 (positive) |
| Test Set |  | 20,845 (negative)<br>2,012 (positive) | 7,196 (negative)<br>384 (positive) | 1,968 (negative)<br>91 (positive) | 207 (negative)<br>29 (positive) |

During the development process, training sets were used for model development, hyperparameter optimization, and training; validation sets were used for model optimization; and after successful development and training, test sets were used to evaluate the performance of the final models. The OUH training set consisted of COVID-free cases prior to the outbreak, and so we matched every COVID-positive case to twenty COVID-free presentations based on age, representing a simulated prevalence of 5%. For PUH, UHB, and BH, we used undifferentiated test sets representing all patients admitted to the respective trusts in the defined time periods.

### Feature Set and Preprocessing

To train and validate our models, we used clinical data with linked, deidentified demographic information for all patients presenting to emergency departments across the four hospital groups. To better compare our results to the clinical validation study performed by Soltan et al. (2022), we used a similar set of features to one of their models – “CURIAL-Lab” – which used a focused subset of routinely collected clinical features. These included blood tests and vital signs, excluding the coagulation panel and blood gas testing, as these are not performed universally and are less informative (Soltan et al., 2022). However, unlike CURIAL-Lab, we did not include the type of oxygen delivery device as a feature, as it is not coded universally between sites (requiring custom preprocessing in order to make the test site data equivalent) and, as neural networks evaluate features heavily on their variability with respect to other variables, we wanted to use a feature set consisting of entirely continuous variables to avoid any optimization or convergence issues. Subsequently, we also did not include oxygen saturation, as this is clinically uninterpretable without knowledge of how much oxygen support is needed. Table 3 summarizes the final features included.

**Table 3:** Clinical predictors considered. (ALT: alanine aminotransferase; CRP: C-reactive protein; eGFR: estimated glomerular filtration rate)

| Category | Features |
| --- | --- |
| Vital Signs | Heart rate, respiratory rate, systolic blood pressure, diastolic blood pressure, temperature |
| Full Blood Count (FBC) | Haemoglobin, haematocrit, mean cell volume, white cell count, neutrophil count, lymphocyte count, monocyte count, eosinophil count, basophil count, platelets |
| Urea and Electrolytes (U&Es) | Sodium, potassium, creatinine, urea, eGFR |
| Liver Function (LF) Tests and CRP | Albumin, alkaline phosphatase, ALT, bilirubin, CRP |

Consistent with Soltan et al. (2022), we used population median imputation to replace any missing values. We then standardized all features in our data to have a mean of 0 and a standard deviation of 1. To ensure that all test sets were treated independently from the training data, on the assumption that the training data are not accessible at the point of modelling the data from the test sites, preprocessing was performed independently on each target site. Thus, imputation and standardization methods based on the training data were used to preprocess the training and validation sets; the test sets were preprocessed using the same methods based instead on each respective test cohort. For example, the standardization requires knowledge of the variance of each variable; in preprocessing the test data, the variance was thus derived from the test data and not from the training data. This ensures that there is no distribution leakage between the training and test sets, allowing for unbiased external evaluation, which is the hypothesis that we seek to test. Additionally, because test sets are preprocessed independently to the training sets, all comparator models are tested on the same test cohorts, allowing for direct comparison of the various methods.

### Model Architecture and Optimization

To predict COVID-19 status, we used a fully-connected neural network (Fig. 1). The rectified linear unit (ReLU) activation function was used for the hidden layers and the sigmoid activation function was used in the output layer. For updating model weights, the Adaptive Moment Estimation (Adam) optimizer was used during training.

**Figure 1:**
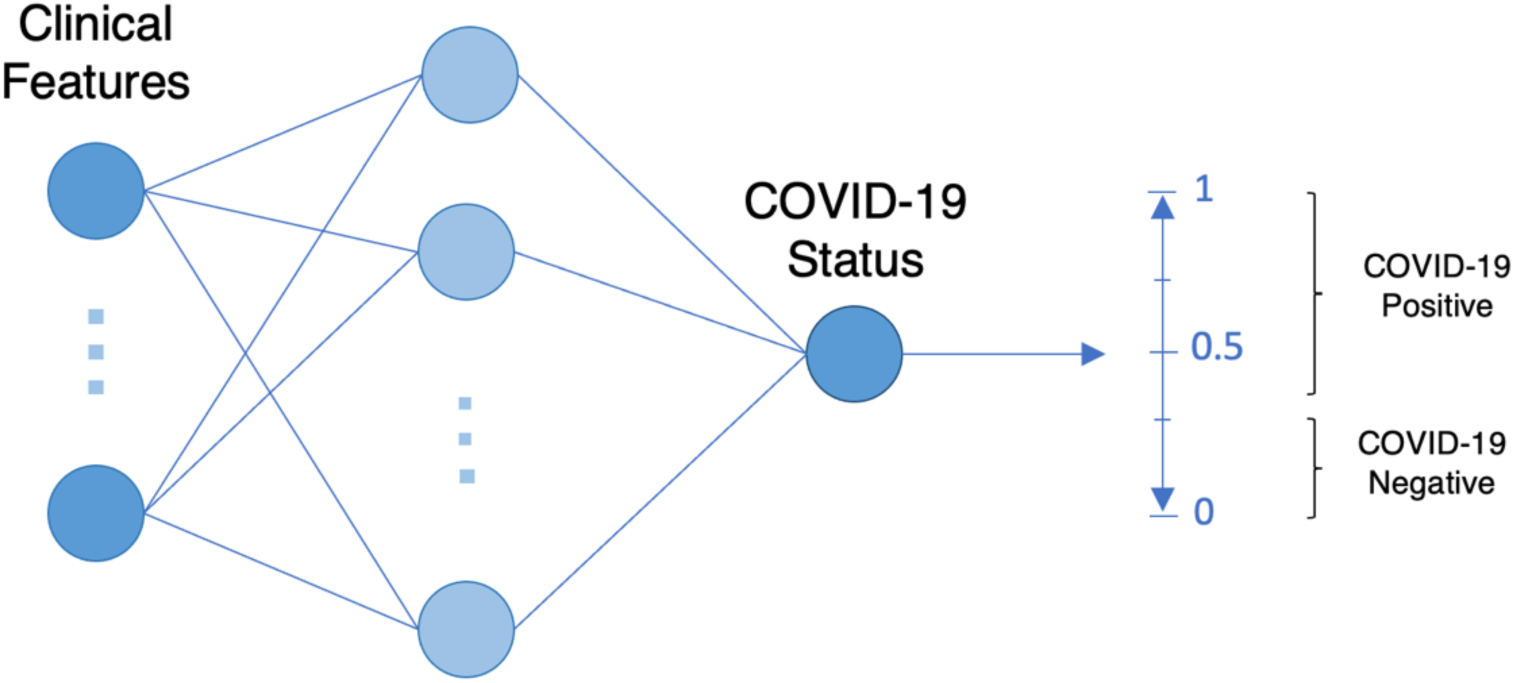
Neural network architecture used for COVID-19 status prediction.

For each model developed, we determined appropriate hyperparameter values through standard 5-fold cross-validation, using respective training sets. We performed a grid search to determine: (i) the number of nodes to be used in each layer of the neural network, (ii) the learning rate, and (iii) the dropout rate. Details on the hyperparameter values used in the final models can be found in Supplementary Table S1.

The raw output of many machine learning classification algorithms is a probability of class membership, which must be interpreted before it’s mapped to a particular class label (Fig. 1). For binary classification, the default threshold is typically 0.5, where all values equal to or greater than 0.5 are mapped to one class and all other values are mapped to the other. However, this default threshold can lead to poor performance, especially when there is a large class imbalance (as seen with our training datasets). Thus, we used a grid search to adjust the decision boundary used for identifying COVID-19 positive or negative cases. For our purposes, we optimized this threshold to sensitivities of 0.85 to ensure clinically acceptable performance in detecting positive COVID-19 cases, and to exceed the sensitivity of lateral flow device (LFD) tests which are used in routine care. LFD sensitivity for OUH admissions between December 23, 2021 and March 6, 2021 was 56.9% (95% confidence interval 51.7-62.0) (Soltan et al., 2022).

### Training Outline

#### Baseline Models

To start, we established baseline models for single-site and multi-site training. For the multi-site baseline, we trained and optimized a model using the combined training data and combined validation data from all the sites. Here, preprocessing was performed after the data had been combined. This model was then tested on all four test sets, separately.

For the single-site baseline models, we used data from each site separately in training, building four custom models. We then subsequently tested each model on the held-out test set from the same site used for training, akin to internal validation.

After training the two baseline models described, we then evaluated three different methods for adopting the single-site models in new settings (for use as ready-made models), akin to external validation. The three methods are: 1) Testing models “as-is”, 2) Readjusting model output thresholds using test site-specific data, and 3) Finetuning models (via transfer learning) using test site-specific data.

#### Testing “as-is”

The first method applies a ready-made model “as-is,” without any modifications. Thus, for the previously trained single-site models, we directly evaluated them on each of the external test sets. This method can be used when the external site does not have access to the original training data, preprocessing transforms, model architecture, and model weights.

#### Threshold adjustment

To adapt a ready-made model to a new setting, a ready-made model can be adjusted, via re-thresholding, using test site-specific data. For each site tested, we used the site-specific validation set to readjust the decision boundary on each ready-made model (i.e., optimizing the model to a sensitivity of 0.85, based on the new site’s data). The readjusted model was then evaluated on its respective test set. This method can also be used when the external site does not have access to the original training data, preprocessing transforms, model architecture, or model weights, as it only modifies the decision threshold of the final model output (recall Fig. 1).

#### Transfer learning

The final method adapts a ready-made model to a new setting using transfer learning. For each site tested, we used the site-specific training and validation set to finetune each ready-made model by updating the existing weights. For each site, we randomly sampled 50% of the training data to perform transfer learning, modelling real-world scenarios where new sites may not have a lot of data to train standalone models. We used a small learning rate (0.0001) so that the procedure would not completely override the previously learned weights as might otherwise occur with a larger value of the learning rate. As before, the test site-specific validation set was used to optimize the model to a sensitivity of 0.85. The resultant finetuned model was then evaluated on its respective test set. This technique can be used when the external site does not have access to the original training data or the preprocessing pipeline (including functions, transforms, etc.), but does have access to the model architecture and pre-trained model weights.

## Results

We evaluated three frameworks for adopting ready-made models to new settings – directly using models “as-is”, readjusting model thresholds using test site-specific data, and finetuning models using test site-specific data via transfer learning. These were compared to two baseline models – a site-specific model and a multi-site trained model. We rigorously processed our data such that test sets from each site were treated independent of the training and validation data, mirroring real-world scenarios where ready-made models are available for others to use, but not the original training data or any of the preprocessing components. Each model was evaluated on the same held-out test sets from each of the four sites and optimized to sensitivities of 0.85. The performance varied between the different testing approaches (Fig. 2); however, all models, including baseline and ready-made approaches, achieved high prevalence-dependent NPV scores (>0.959), demonstrating the ability to exclude a COVID-19 with high-confidence.

**Figure 2:**
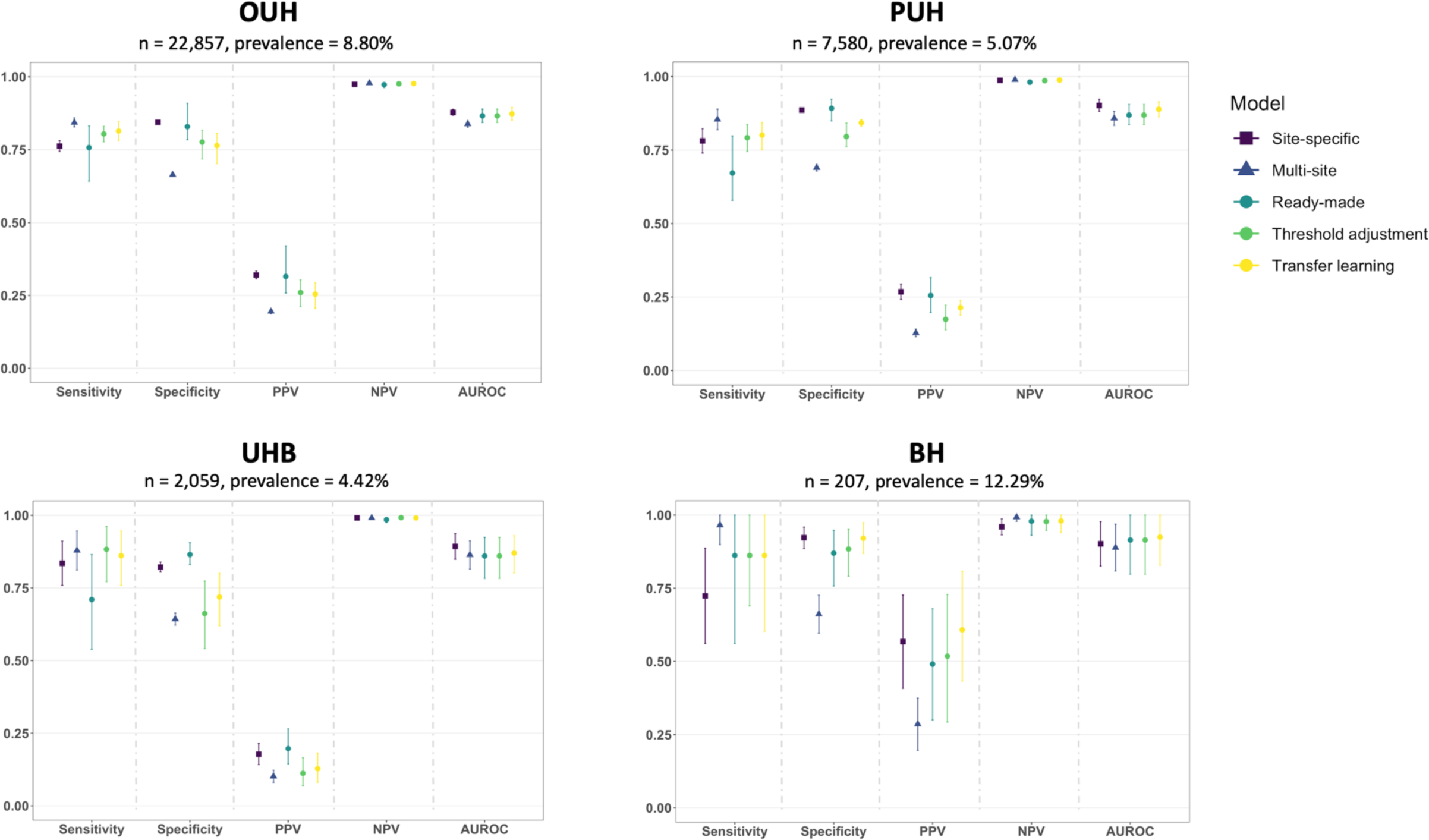
Performance of different model-implementation methods (ready-made, threshold adjustment, transfer learning) and baseline models (site-specific and multi-site), across four NHS Trusts (numerical results are shown in Supplementary Tables S2 and S3). All models were optimized during training and validation to achieve sensitivities of 0.85 (optimal thresholds can be found in Supplementary Table S4). Error bars show 95% confidence intervals (CIs) based on standard error. CIs for AUROC are calculated using Hanley and McNeil’s method.

Of the ready-made model techniques, transfer learning consistently achieved the best results in terms of AUROC, achieving mean AUROCs of 0.873 (95% CIs range 0.851-0.895), 0.889 (0.863-0.914), 0.870 (0.802-0.930), and 0.925 (0.829-1.000) for OUH, PUH, UHB, and BH test sets, respectively. Transfer learning performed on the ready-made model from BH had the lowest performances (AUROCs of 0.862 [95% CI 0.851-0.872], 0.885 [0.863-0.907], and 0.852 [0.802-0.901] when finetuning with OUH, PUH and UHB data, respectively). Overall, transfer learning resulted in better performance than applying both a ready-made model “as-is” and performing threshold adjustment, per training site, individually (p << 0.05 for all site-specific models, Supplementary Table S5). AUROCs achieved by transfer learning were comparable to those set by the baseline site-specific models (AUROCs 0.878 [95% CI 0.868-0.888], 0.902 [0.882-0.923], 0.893 [0.849-0.937], and 0.902 [0.826-0.978] for OUH, PUH, UHB, and BH, respectively). In general, when using ready-made models, the lowest AUROCs were from applying ready-made models “as-is” and applying the models after threshold adjustment. The raw probability output from these two methods are the same, as threshold adjustment occurs after a model is already trained; thus, they had the same AUROC scores (mean AUROCs of 0.866 [95% CIs range 0.843-0.889], 0.869 [0.837-0.905], 0.860 [0.783-0.924], 0.915 [0.798-1.000]). Although the AUROC scores were generally lower than those achieved through transfer learning, both applying ready-made models “as-is” and using threshold adjustment achieved higher mean AUROCs than the multi-site model at all test-sites except UHB, where it achieved a similar score.

In terms of sensitivity, the highest scores tended to be from threshold adjustment (mean sensitivities of 0.804 [95% CIs range 0.777-0.830], 0.792 [0.745-0.837], 0.883 [0.772-0.962], 0.862 [0.690-1.000], for OUH, PUH, UHB, and BH test sets, respectively) and transfer learning (mean sensitivities 0.814 [0.781-0.846], 0.801 [0.751-0.844], 0.861 [0.759-0.946], 0.862 [0.603-1.000]). And, as before, using the models directly “as-is” tended to achieve the lowest performances (mean sensitivities of 0.757 [95% CIs range 0.642-0.831], 0.672 [0.579-0.798], 0.710 [0.539-0.865], 0.862 [0.561-1.000]), as well as exhibited the greatest variability, as reflected by the large CI ranges (Fig. 2).

Overall, models that were trained (at any stage) using data from the test site achieved better results, suggesting that site-specific customization is crucial to predictive performance. Summary tables of all individual results can be found in Supplementary Table S2 and mean results (per test site) can be found in Supplementary Table S3.

## Discussion

In many ML-based healthcare studies, there is difficulty achieving multi-site testing and sharing of code and data, resulting in a gap between locally reported performance and cross-site generalizability. In this study, we investigated three approaches for adopting ready-made ML models for use in new, independent hospital settings (using models “as-is”, threshold adjustment using test site-specific data, and finetuning models using test site-specific data via transfer learning). All models were evaluated using clinical data from four NHS Trusts and compared against two baseline models (site-specific models and a multi-site model).

We found that predictive performance varied across different model-testing approaches, with the best performances (in terms of AUROC) resulting from transfer learning on test site-specific data. This is expected, as neural networks often require a lot of data to train; thus, through transfer learning, knowledge learned from a previous task (in our case, a previous hospital setting) can be leveraged for a new one. Transfer learning performed on the ready-made model from BH had the lowest performances. This may be because the BH dataset had the fewest presentations (1,177 patients total, compared to >10,000 patients in each of the other datasets), and thus may be harder to generalize from. However, compared to the other ready-made approaches, transfer learning achieved better performances (per site-specific model) at all test sites, including BH. Although transfer learning was found to be the most effective method, it has the most prerequisites, as it relies on both the model architecture and pre-trained weights to be made available. Also, as cohort distribution can vary across hospital settings, new patient examples may differ greatly from previous training examples. This could lead to catastrophic forgetting (i.e., the neural network forgetting the old data) when performing transfer learning on these new examples. We tried to address this by using a small learning rate, however, this may be ineffective if the differences in the data are sufficiently significant. Thus, new methods should be explored to deal with this issue, such as freezing select network weights. This would especially be useful when the model being applied is much larger/deeper. Here, we performed transfer learning on the entire network, as it was a shallow, single hidden layer network; however, in the case of a deep neural network, finetuning could be performed on just a select few layers, rather than the entire network.

Adjusting the threshold on a ready-made model is a straightforward method for quickly modifying a model’s sensitivity or specificity threshold, as it does not require access to the preprocessing pipeline, the original training data, the model architecture, or the model weights. Although this method achieved lower AUROC scores, it still performed well in terms of sensitivity, as optimization was based on a sensitivity threshold of 0.85; and thus, ensuring that COVID-19 could be detected. As model weights are not changed in this approach, solely adjusting the threshold may lead to a higher number of false positives (hence, at times, the lower AUROC). However, depending on the clinical question, this outcome may still be acceptable.

Thus, if it is logical to simply maximize either sensitivity or specificity (possibly at the expense of the other), then this method may be appropriate to use when adopting a ready-made model in a new setting. Furthermore, it is desirable for a model to achieve consistent sensitivity/specificity scores across different sites. Even if AUROC is consistent, there can still be varying trade-offs between sensitivity and specificity, making it difficult for clinicians to reliably trust the performance characteristics of the model.

Of the ready-made application techniques, applying ready-made models “as-is” resulted in the lowest mean predictive performance across all test sites, as well as the largest variability between models in terms of sensitivity. This is expected, as different hospital settings can vary in terms of unobserved confounders, protocols, and cohort distribution, making prediction difficult to generalize. Even if human pathophysiology may be similar for a particular outcome, neural networks are heavily dependent on the specific datasets and patient cohorts used during training. However, an advantage of a “ready-made” approach is that it can be deployed immediately in a hospital before exposure to COVID-19, whereas the threshold adjustment and transfer learning methods require an initial handful of COVID-19 cases at the site before the model can be used. Alternatively, a possible deployment pipeline could start by using a ready-made model “as-is”, with gradual adjustment/finetuning over time.

Predictive performances achieved through transfer learning were comparable to those of the baseline site-specific models, even when a smaller set of data was used for training (recall, only 50% of the training data used in the baseline site-specific model was used for transfer learning). This demonstrates that transfer learning is a powerful technique for site-specific customization, even when there is less data available. Thus, in general, ML-based healthcare models would greatly benefit from some level of site-specific customization. However, depending on the clinical question, it is still important to consider whether a site-specific model or a multi-site (more generalized) model is best suited for the task. For example, if the purpose of a model is to support patients within a specific hospital care structure, then a site-specific model may be the most appropriate choice.

## Conclusion

In this study, we introduced three methods for adopting single-site, ready-made ML models for use in new hospital settings. We evaluated these approaches through a case study of COVID-19 diagnosis across four independent NHS Hospital Trusts, providing the first direct comparison of cross-site implementation strategies for ready-made ML-based healthcare models. We found that site-specific customization is crucial to predictive performance, demonstrating how ready-made models exhibited poorer performance when directly applied to new settings without any site-specific corrections. The frameworks we introduced can be adapted to many clinical questions and model architectures. Thus, we hope that the ability to adapt ready-made models will both encourage and improve the application, comparison, and reproducibility of ML-driven insights. Especially when code and training data are not publicly accessible, methods for adopting ready-made models will be key to driving forward evidence-based ML within healthcare.

## Data Availability

Data from OUH studied here are available from the Infections in Oxfordshire Research Database (https://oxfordbrc.nihr.ac.uk/research-themesoverview/antimicrobial-resistance-and-modernising-microbiology/infections-inoxfordshire-research-database-iord/), subject to an application meeting the ethical and governance requirements of the Database. Data from UHB, PUH and BH are available on reasonable request to the respective trusts, subject to HRA requirements. Code and supplementary information for this paper are available online alongside publication.

## Contributions

JY conceived the study. JY & AS designed the study, and accessed, verified, and preprocessed the data. JY performed the analyses and wrote the manuscript. All authors had access to all of the study data and revised the manuscript.

## Acknowledgements

We express our sincere thanks to all patients and staff across the four participating NHS trusts; Oxford University Hospitals NHS Foundation Trust, University Hospitals Birmingham NHS Trust, Bedfordshire Hospitals NHS Foundations Trust, and Portsmouth Hospitals University NHS Trust. We additionally express our gratitude to Jingyi Wang & Dr Jolene Atia at University Hospitals Birmingham NHS Foundation trust, Phillip Dickson at Bedfordshire Hospitals, and Paul Meredith at Portsmouth Hospitals University NHS Trust for assistance with data extraction.

## Ethics

United Kingdom National Health Service (NHS) approval via the national oversight/regulatory body, the Health Research Authority (HRA), has been granted for this work (IRAS ID: 281832).

