## Supplementary Material for "Machine Learning Generalizability Across Healthcare Settings: Insights from multi-site COVID-19 screening"

Jenny Yang et al., 2022

### Appendix A:

Inclusion & Exclusion Criteria:

*Oxford University Hospitals NHS Foundation Trust (OUH):*

We included all patients attending acute and emergency care settings at OUH who received routine blood tests on arrival, considering presentations before December 1, 2019, and thus before the pandemic, as the COVID-19-negative (control) cohort. We considered presentations during the 'first wave' of the UK COVID-19 pandemic (December 1, 2019 to June 30, 2020) with PCR confirmed SARS-CoV-2 infection as the COVID-19-positive (cases) cohort. We excluded patients who opted out of electronic health record (EHR) research and those who did not receive laboratory blood tests or were younger than 18 years of age. Due to incomplete penetrance of testing during the first wave of the pandemic, and imperfect sensitivity of the PCR test, there is uncertainty in the viral status of patients presenting during the pandemic who were untested or tested negative. We therefore selected a pre-pandemic control cohort during training to ensure absence of disease in patients labelled as COVID-19-negative. Clinical features extracted for each presentation included first-performed blood tests, blood gases, vital signs measurements and PCR testing for SARS-CoV-2 (Abbott Architect [Abbott, Maidenhead, UK], TaqPath [Thermo Fisher Scientific, Massachusetts, USA] and Public Health England-designed RNA-dependent RNA polymerase assays).

*Portsmouth Hospitals NHS Foundation Trust (PUH):*

PUH considered all patients admitted to the Queen Alexandria Hospital, serving a population of 675,000 and offering tertiary referral services to the surrounding region, between March 1, 2020 and February 28, 2021. Confirmatory COVID-19 testing was by laboratory SARS-CoV2 RT-PCR assay, considering any positive PCR result within 48hrs of admission as a true positive.

*University Hospitals Birmingham NHS Foundation Trust (UHB):*

UHB considered all patients admitted to The Queen Elizabeth Hospital, Birmingham, between December 01, 2019 and October 29, 2020. The Queen Elizabeth Hospital is a large tertiary referral unit within the UHB group which provides healthcare services for a population of 2.2 million across the West Midlands. Confirmatory COVID-19 testing was performed by laboratory SARS-CoV-2 RT-PCR assay.

*Bedfordshire NHS Foundation Trust (BH):*

BH considered all patients admitted to Bedford Hospital between January 1, 2021 and March 31, 2021. BH provides healthcare services for a population of around 620,000 in Bedfordshire. Confirmatory COVID-19 testing was performed on the day of admission by point-of-care PCR based nucleic acid testing [SAMBA-II & Panther Fusion System, Diagnostics in the Real World, UK, and Hologic, USA].

### Appendix B:

Table S1: Final hyperparameter values used in models.

| Model | Nodes | Dropout Rate | Learning Rate | Epochs | Batch Size |
|---|---|---|---|---|---|
| OUH | 10 | 0.3 | 0.1 | 30 | 10 |
| PUH | 20 | 0.3 | 0.01 | 30 | 10 |
| UHB | 10 | 0.5 | 0.01 | 30 | 10 |
| BH | 10 | 0.3 | 0.01 | 30 | 10 |
| Multi-site | 10 | 0.5 | 0.01 | 30 | 10 |

Table S2: Performance results of different models. Results are reported alongside 95% confidence intervals (CIs) based on standard error. CIs for AUROC are calculated using Hanley and McNeil's method.

| Validation Site | Model | Sensitivity | Specificity | PPV | NPV | F1 | AUROC |
|---|---|---|---|---|---|---|---|
| OUH | Site-specific | 0.762 (0.744-0.781) | 0.844 (0.839-0.849) | 0.320 (0.307-0.333) | 0.974 (0.971-0.976) | 0.451 | 0.878 (0.868-0.888) |
| | Multi-site | 0.843 (0.828-0.859) | 0.664 (0.658-0.671) | 0.195 (0.187-0.203) | 0.978 (0.975-0.980) | 0.317 | 0.838 (0.827-0.849) |
| | Ready-made (PUH) | 0.814 (0.797-0.831) | 0.790 (0.784-0.795) | 0.272 (0.261-0.283) | 0.978 (0.976-0.980) | 0.408 | 0.879 (0.869-0.889) |
| | Ready-made (UHB) | 0.795 (0.777-0.812) | 0.792 (0.786-0.797) | 0.269 (0.258-0.280) | 0.976 (0.973-0.978) | 0.402 | 0.867 (0.857-0.877) |
| | Ready-made (BH) | 0.663 (0.642-0.683) | 0.905 (0.901-0.909) | 0.403 (0.387-0.420) | 0.965 (0.963-0.968) | 0.502 | 0.853 (0.843-0.864) |
| | Threshold Adjustment (PUH) | 0.804 (0.787-0.822) | 0.811 (0.806-0.816) | 0.291 (0.279-0.303) | 0.977 (0.975-0.979) | 0.427 | 0.879 (0.869-0.889) |
| | Threshold Adjustment (UHB) | 0.795 (0.777-0.812) | 0.792 (0.786-0.797) | 0.269 (0.258-0.280) | 0.976 (0.973-0.978) | 0.402 | 0.867 (0.857-0.877) |
| | Threshold Adjustment (BH) | 0.813 (0.796-0.830) | 0.724 (0.718-0.730) | 0.221 (0.212-0.231) | 0.976 (0.973-0.978) | 0.348 | 0.853 (0.843-0.864) |
| | Transfer learning (PUH) | 0.813 (0.796-0.830) | 0.800 (0.795-0.806) | 0.282 (0.270-0.294) | 0.978 (0.976-0.980) | 0.419 | 0.885 (0.875-0.895) |
| | Transfer learning (UHB) | 0.799 (0.781-0.816) | 0.785 (0.779-0.791) | 0.264 (0.253-0.275) | 0.976 (0.974-0.978) | 0.397 | 0.872 (0.862-0.882) |
| | Transfer learning (BH) | 0.830 (0.814-0.846) | 0.708 (0.702-0.714) | 0.215 (0.206-0.225) | 0.977 (0.975-0.980) | 0.342 | 0.862 (0.851-0.872) |
| PUH | Site-specific | 0.781 (0.740-0.823) | 0.886 (0.879-0.893) | 0.268 (0.242-0.294) | 0.987 (0.984-0.990) | 0.399 | 0.902 (0.882-0.923) |
| | Multi-site | 0.854 (0.819-0.889) | 0.690 (0.679-0.701) | 0.128 (0.115-0.141) | 0.989 (0.986-0.992) | 0.223 | 0.858 (0.834-0.882) |
| | Ready-made (OUH) | 0.633 (0.585-0.681) | 0.903 (0.897-0.910) | 0.259 (0.231-0.287) | 0.979 (0.975-0.982) | 0.368 | 0.861 (0.837-0.885) |
| | Ready-made (UHB) | 0.755 (0.712-0.798) | 0.857 (0.849-0.865) | 0.220 (0.198-0.243) | 0.985 (0.982-0.988) | 0.341 | 0.883 (0.861-0.905) |
| | Ready-made (BH) | 0.628 (0.579-0.676) | 0.916 (0.910-0.923) | 0.286 (0.255-0.316) | 0.979 (0.975-0.982) | 0.393 | 0.862 (0.838-0.885) |
| | Threshold Adjustment (OUH) | 0.797 (0.757-0.837) | 0.784 (0.774-0.793) | 0.164 (0.147-0.181) | 0.986 (0.983-0.989) | 0.272 | 0.861 (0.837-0.885) |
| | Threshold Adjustment (UHB) | 0.792 (0.751-0.832) | 0.833 (0.825-0.842) | 0.202 (0.182-0.222) | 0.987 (0.984-0.990) | 0.322 | 0.883 (0.861-0.905) |
| | Threshold Adjustment (BH) | 0.786 (0.745-0.827) | 0.771 (0.761-0.781) | 0.155 (0.139-0.171) | 0.985 (0.982-0.989) | 0.259 | 0.862 (0.838-0.885) |
| | Transfer learning (OUH) | 0.805 (0.765-0.844) | 0.845 (0.837-0.854) | 0.217 (0.196-0.239) | 0.988 (0.985-0.991) | 0.342 | 0.890 (0.868-0.911) |
| | Transfer learning (UHB) | 0.805 (0.765-0.844) | 0.845 (0.837-0.853) | 0.217 (0.196-0.238) | 0.988 (0.985-0.991) | 0.342 | 0.892 (0.871-0.914) |
| | Transfer learning (BH) | 0.792 (0.751-0.832) | 0.839 (0.831-0.848) | 0.208 (0.188-0.229) | 0.987 (0.984-0.990) | 0.330 | 0.885 (0.863-0.907) |
| UHB | Site-specific | 0.835 (0.759-0.911) | 0.822 (0.805-0.839) | 0.178 (0.142-0.215) | 0.991 (0.986-0.995) | 0.294 | 0.893 (0.849-0.937) |
| | Multi-site | 0.879 (0.812-0.946) | 0.643 (0.622-0.664) | 0.102 (0.081-0.123) | 0.991 (0.986-0.996) | 0.183 | 0.864 (0.815-0.912) |
| | Ready-made (OUH) | 0.714 (0.621-0.807) | 0.854 (0.839-0.870) | 0.185 (0.144-0.225) | 0.985 (0.979-0.991) | 0.293 | 0.878 (0.832-0.924) |
| | Ready-made (PUH) | 0.780 (0.695-0.865) | 0.847 (0.831-0.862) | 0.190 (0.151-0.230) | 0.988 (0.983-0.993) | 0.306 | 0.866 (0.818-0.914) |
| | Ready-made (BH) | 0.637 (0.539-0.736) | 0.893 (0.879-0.906) | 0.216 (0.166-0.265) | 0.982 (0.975-0.988) | 0.322 | 0.835 (0.783-0.887) |
| | Threshold Adjustment (OUH) | 0.901 (0.840-0.962) | 0.669 (0.648-0.690) | 0.112 (0.089-0.135) | 0.993 (0.989-0.998) | 0.199 | 0.878 (0.832-0.924) |
| | Threshold Adjustment (PUH) | 0.846 (0.772-0.920) | 0.755 (0.736-0.774) | 0.137 (0.109-0.166) | 0.991 (0.986-0.996) | 0.237 | 0.866 (0.818-0.914) |
| | Threshold Adjustment (BH) | 0.901 (0.840-0.962) | 0.563 (0.541-0.585) | 0.087 (0.069-0.105) | 0.992 (0.987-0.997) | 0.159 | 0.835 (0.783-0.887) |
| | Transfer learning (OUH) | 0.868 (0.799-0.938) | 0.735 (0.715-0.754) | 0.131 (0.104-0.158) | 0.992 (0.987-0.996) | 0.228 | 0.885 (0.840-0.930) |
| | Transfer learning (PUH) | 0.835 (0.759-0.911) | 0.782 (0.764-0.800) | 0.150 (0.119-0.182) | 0.990 (0.985-0.995) | 0.255 | 0.874 (0.827-0.921) |
| | Transfer learning (BH) | 0.879 (0.812-0.946) | 0.641 (0.620-0.662) | 0.102 (0.081-0.123) | 0.991 (0.986-0.996) | 0.182 | 0.852 (0.802-0.901) |
| BH | Site-specific | 0.724 (0.561-0.887) | 0.923 (0.886-0.959) | 0.568 (0.408-0.727) | 0.960 (0.933-0.987) | 0.636 | 0.902 (0.826-0.978) |
| | Multi-site | 0.966 (0.899-1.000) | 0.662 (0.597-0.726) | 0.286 (0.196-0.375) | 0.993 (0.979-1.000) | 0.441 | 0.889 (0.809-0.969) |
| | Ready-made (OUH) | 0.724 (0.561-0.887) | 0.908 (0.869-0.948) | 0.525 (0.370-0.680) | 0.959 (0.931-0.987) | 0.609 | 0.880 (0.798-0.963) |
| | Ready-made (PUH) | 0.966 (0.899-1.000) | 0.812 (0.758-0.865) | 0.418 (0.300-0.536) | 0.994 (0.983-1.000) | 0.583 | 0.940 (0.880-1.000) |
| | Ready-made (UHB) | 0.897 (0.786-1.000) | 0.889 (0.846-0.932) | 0.531 (0.391-0.670) | 0.984 (0.966-1.000) | 0.667 | 0.924 (0.856-0.992) |
| | Threshold Adjustment (OUH) | 0.828 (0.690-0.965) | 0.841 (0.791-0.890) | 0.421 (0.293-0.549) | 0.972 (0.948-0.996) | 0.558 | 0.880 (0.798-0.963) |
| | Threshold Adjustment (PUH) | 0.862 (0.737-0.988) | 0.913 (0.875-0.951) | 0.581 (0.434-0.729) | 0.979 (0.959-0.999) | 0.694 | 0.940 (0.880-1.000) |
| | Threshold Adjustment (UHB) | 0.897 (0.786-1.000) | 0.899 (0.857-0.940) | 0.553 (0.411-0.695) | 0.984 (0.966-1.000) | 0.684 | 0.924 (0.856-0.992) |
| | Transfer learning (OUH) | 0.759 (0.603-0.914) | 0.942 (0.910-0.974) | 0.647 (0.486-0.808) | 0.965 (0.940-0.991) | 0.698 | 0.904 (0.829-0.979) |
| | Transfer learning (PUH) | 0.897 (0.786-1.000) | 0.908 (0.869-0.948) | 0.578 (0.433-0.722) | 0.984 (0.967-1.000) | 0.703 | 0.944 (0.885-1.000) |
| | Transfer learning (UHB) | 0.931 (0.839-1.000) | 0.913 (0.875-0.951) | 0.600 (0.457-0.743) | 0.990 (0.975-1.000) | 0.730 | 0.928 (0.862-0.994) |

Table S3: Mean results across ready-made models. Results are reported alongside 95% confidence intervals (CIs) based on standard error. CIs for AUROC are calculated using Hanley and McNeil's method.

| Validation Site | Model | Sensitivity | Specificity | PPV | NPV | AUROC |
|---|---|---|---|---|---|---|
| OUH | Ready-made | 0.757 (0.642-0.831) | 0.829 (0.784-0.909) | 0.315 (0.258-0.42) | 0.973 (0.963-0.980) | 0.866 (0.843-0.889) |
| | Threshold Adjustment | 0.804 (0.777-0.830) | 0.776 (0.718-0.816) | 0.260 (0.212-0.303) | 0.976 (0.973-0.979) | 0.866 (0.843-0.889) |
| | Transfer learning | 0.814 (0.781-0.846) | 0.764 (0.702-0.806) | 0.254 (0.206-0.294) | 0.977 (0.974-0.980) | 0.873 (0.851-0.895) |
| PUH | Ready-made | 0.672 (0.579-0.798) | 0.892 (0.849-0.923) | 0.255 (0.198-0.316) | 0.981 (0.975-0.988) | 0.869 (0.837-0.905) |
| | Threshold Adjustment | 0.792 (0.745-0.837) | 0.796 (0.761-0.842) | 0.174 (0.139-0.222) | 0.986 (0.982-0.990) | 0.869 (0.837-0.905) |
| | Transfer learning | 0.801 (0.751-0.844) | 0.843 (0.831-0.854) | 0.214 (0.188-0.239) | 0.988 (0.984-0.991) | 0.889 (0.863-0.914) |
| UHB | Ready-made | 0.710 (0.539-0.865) | 0.865 (0.831-0.906) | 0.197 (0.144-0.265) | 0.985 (0.975-0.993) | 0.860 (0.783-0.924) |
| | Threshold Adjustment | 0.883 (0.772-0.962) | 0.662 (0.541-0.774) | 0.112 (0.069-0.166) | 0.992 (0.986-0.998) | 0.860 (0.783-0.924) |
| | Transfer learning | 0.861 (0.759-0.946) | 0.719 (0.620-0.800) | 0.128 (0.081-0.182) | 0.991 (0.985-0.996) | 0.870 (0.802-0.930) |
| BH | Ready-made | 0.862 (0.561-1.000) | 0.870 (0.758-0.948) | 0.491 (0.300-0.680) | 0.979 (0.931-1.000) | 0.915 (0.798-1.000) |
| | Threshold Adjustment | 0.862 (0.690-1.000) | 0.884 (0.791-0.951) | 0.518 (0.293-0.729) | 0.978 (0.948-1.000) | 0.915 (0.798-1.000) |
| | Transfer learning | 0.862 (0.603-1.000) | 0.921 (0.869-0.974) | 0.608 (0.433-0.808) | 0.980 (0.940-1.000) | 0.925 (0.829-1.000) |

Table S4: Optimization thresholds used during model evaluation.

| **Validation Site** | **Model** | **Optimization Threshold** |
|---|---|---|
| OUH | Site-specific: | 0.052 |
| | Multi-site: | 0.042 |
| | Ready-made (PUH): | 0.064 |
| | Ready-made (UHB): | 0.046 |
| | Ready-made (BH): | 0.200 |
| | Threshold Adjustment (PUH): | 0.074 |
| | Threshold Adjustment (UHB): | 0.046 |
| | Threshold Adjustment (BH): | 0.060 |
| | Transfer learning (PUH): | 0.052 |
| | Transfer learning (UHB): | 0.054 |
| | Transfer learning (BH): | 0.032 |
| PUH | Site-specific: | 0.064 |
| | Multi-site: | 0.042 |
| | Ready-made (OUH): | 0.052 |
| | Ready-made (UHB): | 0.046 |
| | Ready-made (BH): | 0.200 |
| | Threshold Adjustment (OUH): | 0.024 |
| | Threshold Adjustment (UHB): | 0.040 |
| | Threshold Adjustment (BH): | 0.074 |
| | Transfer learning (OUH): | 0.056 |
| | Transfer learning (UHB): | 0.058 |
| | Transfer learning (BH): | 0.056 |
| UHB | Site-specific: | 0.046 |
| | Multi-site: | 0.042 |
| | Ready-made (OUH): | 0.052 |
| | Ready-made (PUH): | 0.064 |
| | Ready-made (BH): | 0.200 |
| | Threshold Adjustment (OUH): | 0.018 |
| | Threshold Adjustment (PUH): | 0.034 |
| | Threshold Adjustment (BH): | 0.022 |
| | Transfer learning (OUH): | 0.030 |
| | Transfer learning (PUH): | 0.034 |
| | Transfer learning (BH): | 0.016 |
| BH | Site-specific: | 0.200 |
| | Multi-site: | 0.042 |
| | Ready-made (OUH): | 0.052 |
| | Ready-made (PUH): | 0.064 |
| | Ready-made (UHB): | 0.046 |
| | Threshold Adjustment (OUH): | 0.030 |
| | Threshold Adjustment (PUH): | 0.156 |
| | Threshold Adjustment (UHB): | 0.050 |
| | Transfer learning (OUH): | 0.082 |
| | Transfer learning (PUH): | 0.134 |
| | Transfer learning (UHB): | 0.066 |

Table S5: Comparison of model outputs (raw probabilities) of ready-made models "as-is" to outputs of the transfer learning models, per training site. The raw probability output of the threshold adjusted models are the same as those from the ready-made models "as-is", as threshold adjustment occurs after a model is already trained. P-values are calculated using the Wilcoxon Signed Rank Test, which was implemented using the statistics package from the SciPy library (numbers < e-308 cannot be distinguished from 0.0 by floating-point types in Python).

| Validation Site | Training Site | p-value |
|---|---|---|
| OUH | PUH | 1.27e-204 |
| | UHB | < e-308 |
| | BH | < e-308 |
| PUH | OUH | < e-308 |
| | UHB | < e-308 |
| | BH | < e-308 |
| UHB | OUH | 2.31e-32 |
| | PUH | 9.83e-7 |
| | BH | 1.19e-202 |
| BH | OUH | 3.02e-29 |
| | PUH | 7.76e-31 |
| | UHB | 1.62e-37 |
